# Detection of SARS-CoV-2 in wastewater in Japan by multiple molecular assays-implication for wastewater-based epidemiology (WBE)

**DOI:** 10.1101/2020.06.09.20126417

**Authors:** Akihiko Hata, Ryo Honda, Hiroe Hara-Yamamura, Yuno Meuchi

## Abstract

Presence of SARS-coronavirus-2 (SARS-CoV-2) in wastewater sample has been documented in several countries. Wastewater-based epidemiology (WBE) is potentially effective for early warning of COVID-19 outbreak. The purpose of this study was to verify the detection limit of WBE for COVID-19. In total, 27 influent wastewater samples were collected from four wastewater treatment plants in Ishikawa and Toyama prefectures in Japan. During the study period, numbers of the confirmed COVID-19 cases in these prefectures increased from almost 0 to around 20 per 100,000 peoples. SARS-CoV-2 RNA in the samples were identified by several PCR-based assays. Among the 27 samples, 7 were positive for SARS-CoV-2 by at least one out of the three quantitative RT-PCR assays. These samples were also positive by RT-nested PCR assays. The detection frequency became higher when the number of total confirmed SARS-CoV-2 cases in 100,000 peoples became above 10 in each prefecture. However, SARS-CoV-2 could also be detected with a low frequency when the number was below 1.0. Considering that the number of the confirmed cases does not necessarily reflect the actual prevalence of the infection at the time point, data on the relationship between the number of infection cases and concentration in wastewater needs to be accumulated further.

## 1. INTRODUCTION

The recent novel coronavirus pneumonia (COVID-19) pandemic caused by the severe acute respiratory syndrome coronavirus 2 (SARS-CoV-2) infection has led to more than 6.8 million of the confirmed cases and nearly 400,000 deaths worldwide, as of June 7, 2020 (WHO, 2020). Currently, this pandemic is likely to be heading for convergence due to world-wide stay-at-home order. However, there are still a concern of re-outbreak after lift of the stay-at-home order for reopening businesses, and also the 2nd pandemic caused by seasonal factors. Wastewater-based epidemiology (WBE) is the effective approach to provide a snapshot of the outbreak situation in the entire catchment by monitoring pathogens and viruses in wastewater (Choi et al., 2018; Yang et al., 2015). The WBE could be an early warning tool for the possible re-outbreaks and seasonal pandemics in the future.

Recently, detection of SARS-CoV-2 in wastewater is reported in Netherlands, Australia, US, China, France, Israel, Italy, Spain, and Japan (Ahmed et al., 2020; Bar-or et al., 2020; Haramoto et al., 2020; La Rosa et al., 2020; Medema et al., 2020; Nemudryi et al., 2020; Randazzo et al., 2020; Rimoldi et al., 2020; WU et al., 2020; Wurtzer et al., 2020a). SARS-CoV-2 are present in wastewater because they are shed from not only in respiratory secretion but also feces of patients (Tang et al., 2020; Wölfel et al., 2020a; J. Zhang et al., 2020; Y. Zhang et al., 2020). Importantly, pre-symptomatic patients and asymptomatic patients also had a viral load in feces as well as symptomatic patients (Tang et al., 2020; Wölfel et al., 2020a; Y. Zhang et al., 2020). Ratio of asymptomatic infection is estimated as 18% (16-20% as 95%-CI) from the cruise ship passengers in Yokohama (Mizumoto et al., 2020), and 31% (7.7-54% as 95%-CI) from the passengers of evacuation flight from Wuhan to Tokyo (Nishiura et al., 2020). Moreover, a much larger population of asymptomatic and mildly symptomatic infections was implied by the surveillance of antibodies to SARS-CoV-2 in Santa Clara County, California, in which the estimated number of infection is 50- to 85-fold more than the number of confirmed cases (Bendavid et al., 2020). However, these people with no symptoms and mild symptoms may not be included in clinical surveillance because most of them do not visit clinics or hospitals. Transmission from such asymptomatic or pre-symptomatic patients makes it difficult to enable the initial containment of COVID-19. Moreover, the number of the reported COVID-19 cases is possibly biased by circumstances of medical services in each country, e.g. access to medical services, capacity of PCR tests, stay-at-home policy for mildly symptomatic patients, situation of medical collapse, etc. So far, Japan has unique trend of COVID-19 epidemic. The increasing trend of cases is much slower than other countries in Europe, US and China (Gloeckner et al., 2020).

Some paper argues that the number of cases is underestimated in Japan because of low capacity of PCR tests (Omori et al., 2020). WBE is able to cover people with asymptomatic and pre-symptomatic infections and to predict the overall epidemic status without such biases of medical situations. Therefore, WBE could be more sensitive and possibly detect outbreak earlier than clinical surveillance and also effective to verify coverage of clinical surveillance under various medical situations. For application of WBE as an early warning tool of COVID-19 outbreak, it is necessary to verify correlations of SARS-CoV-2 concentration in wastewater and the number of COVID-19 cases in each country.

According the recent studies, SARS-CoV-2 were detected in wastewater when the number of confirmed cases reached 1-100 cases per 100,000 of population (Ahmed et al., 2020; Bar-or et al., 2020; Medema et al., 2020; Nemudryi et al., 2020; WU et al., 2020; Wurtzer et al., 2020a). The sensitivity of WBE depends on viral load in feces of infected people. Detection of SARS-CoV-2 in wastewater could be less sensitive than norovirus, because the viral load of SARS-CoV-2 in feces is reportedly 1 to 2 logs lower than norovirus (Hata and Honda, 2020). SARS-CoV-2 is an enveloped virus, while typical viral pathogens in water, such as noroviruses, are non-enveloped viruses. Then, commonly used methods for concentrating viruses in water might not be effective to SARS-CoV-2 (Kitajima et al., 2020). Besides, several RT-PCR assays for SARS-CoV-2 are now available but their applicability to wastewater sample has not been well determined. These indicate that detection of SARS-CoV-2 in the wastewater is more challenging than that of norovirus and other enteric viruses. At this time, verifying concentration methods and downstream RT-PCR assays is necessary to make WBE reliable.

The objective of this study is to verify the detection limit of WBE for COVID-19 by comparing the detected concentration of SARS-CoV-2 in wastewater and the COVID-19 cases reported in clinical surveillance. In this study, wastewater samples were taken in two prefectures, Ishikawa and Toyama, in Japan which had the most reported COVID-19 cases per population except Tokyo Metropolitan. To deal with the methodological issues pointed above, sample processing was validated by assessing virus detection efficiency with process controls. Besides, multiple RT-PCR assays and nucleotide sequencing were applied to improve the reliability of the positive results. At the beginning of the study period, numbers of the confirmed COVID-19 cases in Ishikawa and Toyama prefectures were only 0.3 and 0 per 100,000 peoples (4 and no confirmed cases), respectively. This enabled us to discuss sensitivity of WBE for COVID-19 from the reported COVID-19 cases per population in the target area.

## 2. MATERIALS AND METHODS

### 2.1 Sample collection

Influent wastewater samples were collected at four WWTPs, WWTPs A-C and D, in Ishikawa and Toyama prefecture, respectively. These WWTPs were built to treat maximum volumes of 12,800, 53,300, 156,000, and 82,500 m^3^/day of wastewater in total and are serving population equivalents of 31,510, 112,396, 151,937, and 169,400, respectively (Table 1). At each WWTP, 100 mL of a grab sample was collected in the morning, during the hour of peak flow, weekly or biweekly from March 5 to April 23, 2020. In total, 27 samples were obtained. The samples were kept cool or frozen until the analysis, which took place within 3 days after the collection. The sample was split into 80 mL and 20 mL of subsamples, which were then subjected to virus concentration process and quantification of F-phages, respectively (Fig. 1). At the beginning of the study period, there were only four and zero COVID-19 confirmed cases in Ishikawa and Toyama prefectures, respectively. However, the numbers increased during the period. At the end of April, Ishikawa and Toyama prefectures were ranked as second and third, respectively, in Japan in the number of infections per 100,000 peoples (21.9 and 18.1 cases, respectively) (Fig. 2).

**TABLE 1.** Characteristics of WWTPs studied in this study.

| WWTP | Prefecture | Population served by WWTP | Sewered population rate (%) | Maximum treatment capacity (m<sup>3</sup>/d) | Actual treatment capacity (m<sup>3</sup>/d) |
| --- | --- | --- | --- | --- | --- |
| A | Ishikawa | 31,510 | 91.0 | 12,800 | 9,366 |
| B | Ishikawa | 112,396 | 91.4 | 53,300 | 37,106 |
| C | Ishikawa | 151,937 | 97.2 | 156,000 | 86,446 |
| D | Toyama | 169,400 | 91.1 | 82,500 | 55,639 |

**Figure 1.**
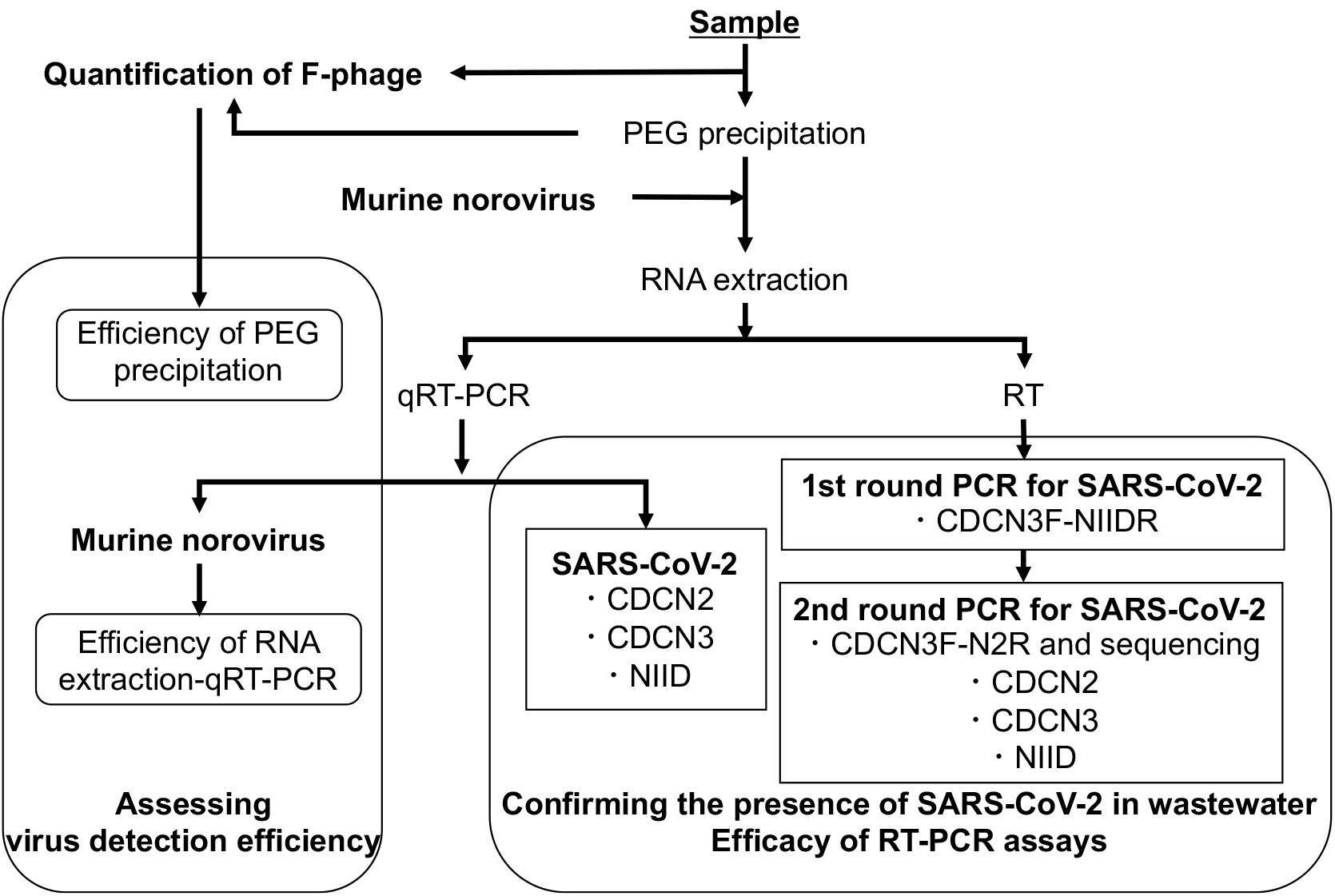
Flow diagram of sample processing for confirming the presence of SARS-CoV-2 in wastewater by RT-PCR assays and for assessing virus detection efficiency with process controls.

**Figure 2.**
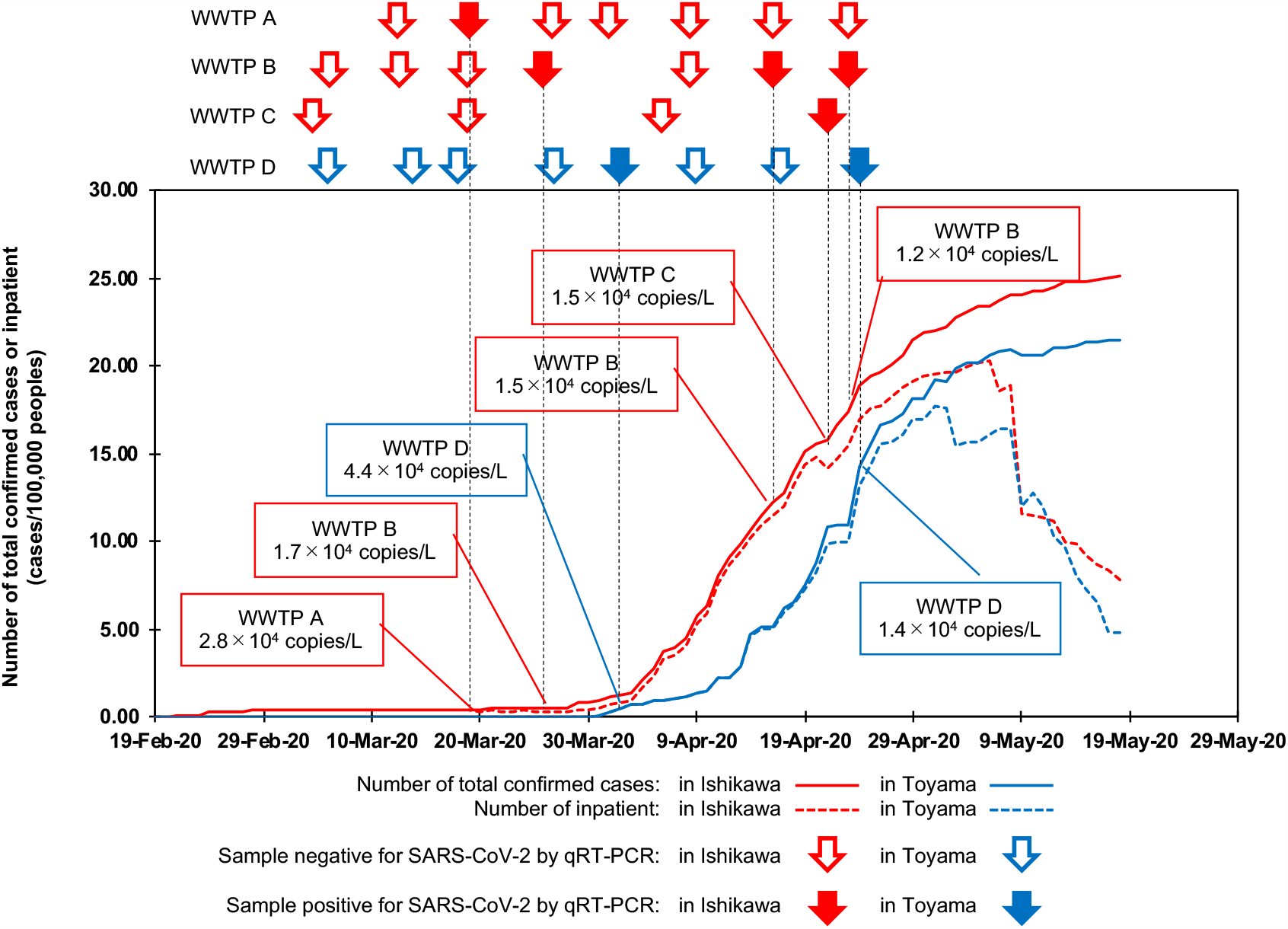
Temporal relationship between number of SARS-CoV-2 infection cases and presence of SARS-CoV-2 in wastewater samples in Ishikawa and Toyama prefectures. Number of inpatients was calculated by subtracting numbers of discharged and dead from number of total confirmed cases, which were derived from a databased summarized by Ministry of Health, Labor and Welfare in Japan (https://www.mhlw.go.jp/stf/seisakunitsuite/bunya/0000121431_00086.html).

### 2.2 Virus concentration by polyethylene glycol precipitation (PEG precipitation)

Prior to the virus concentration, the 80 mL subsample was centrifuged at 5,000×g for 5 minutes. The supernatant was recovered and mixed with 8.0 g of PEG8000 and 4.7 g of sodium chloride to be final concentrations of 10% and 1 M, respectively. The mixture was incubated overnight on a shaker at 4°C. Subsequently, the mixture was centrifuged at 10,000×g for 30 minutes. The supernatant was discarded and the resultant pellet was resuspended in 500 μL of phosphate buffer as a virus concentrate.

### 2.3 RNA extraction

Murine norovirus was used as a molecular process control to monitor the inhibitory effects on the RNA extraction-RT-qPCR processes. Briefly, 140 µL of the virus concentrate was spiked with 1.7×10^4^ copies of MNV and subjected to RNA extraction using a QIAamp viral RNA mini kit (Qiagen) to obtain a 60 µL of RNA extract, in accordance with the manufacturer’s instructions. The obtained RNA extract was subjected to qRT-PCR assays and RT-semi nested PCR assays, for quantitative detection of the spiked MNV and indigenous SARS-CoV-2 and for qualitative detection of indigenous SARS-CoV-2, respectively.

### 2.4 qRT-PCR assay

TaqMan-based qRT-PCR assays for quantitative detection of spiked MNV and indigenous SARS-CoV-2 were performed with a qTOWER^3^ (Analytik Jena). For quantification of MNV RNA, a primer and TaqMan probe set described by Kitajima et al. (2008) was utilized, while for quantification of SARS-CoV-2, two and one primer and TaqMan probe sets described by Centers for Disease Control and Prevention (CDC) in the USA (2020) (CDCN2, and CDCN3 assays, which were originally denoted as 2019-nCoV_N1, 2019-nCoV_N2, and 2019-nCoV_N3, respectively) and Shirato et al. (2020) (NIID assay, which was originally denoted as NIID_2019-nCoV_N), respectively, were separately used (Table 2). Shirato et al. (2020) described two reverse primers for NIID assay. In this study, one of them (NIID_2019-nCoV_N_R2ver3, denoted as NIID-R in Table 2) was used because it does not have mismatch with reference strains (Wuhan-Hu-1 strain, Gen Bank acc. No. of MN908947.3, for example). A reaction mixture (20 μL) was prepared using a One Step PrimeScript™ RT-PCR Kit (Perfect Real Time) (TaKaRa). Briefly, 20 μL of a reaction mixture was prepared by mixing 2 μL or 5 μL of RNA extract with, of 10 μL a 2×One Step RT-PCR Buffer III, 0.4 μL of a TaKaRa Ex Taq HS, 0.4 μL of a PrimeScript RT enzyme Mix II, 400 nM each of forward- and reverse primers, 100 nM of a TaqMan probe, and nuclease-free water. qRT-PCR was performed under the following thermal cycling conditions: RT reaction at 42°C for 5 min, initial denaturation and inactivation of the RT enzyme at 95°C for 10 s, followed by 50 cycles of amplification with denaturation at 95°C for 5 s and annealing and extension at specific temperatures for each assay (Table 2) for 30 s.

**TABLE 2.**
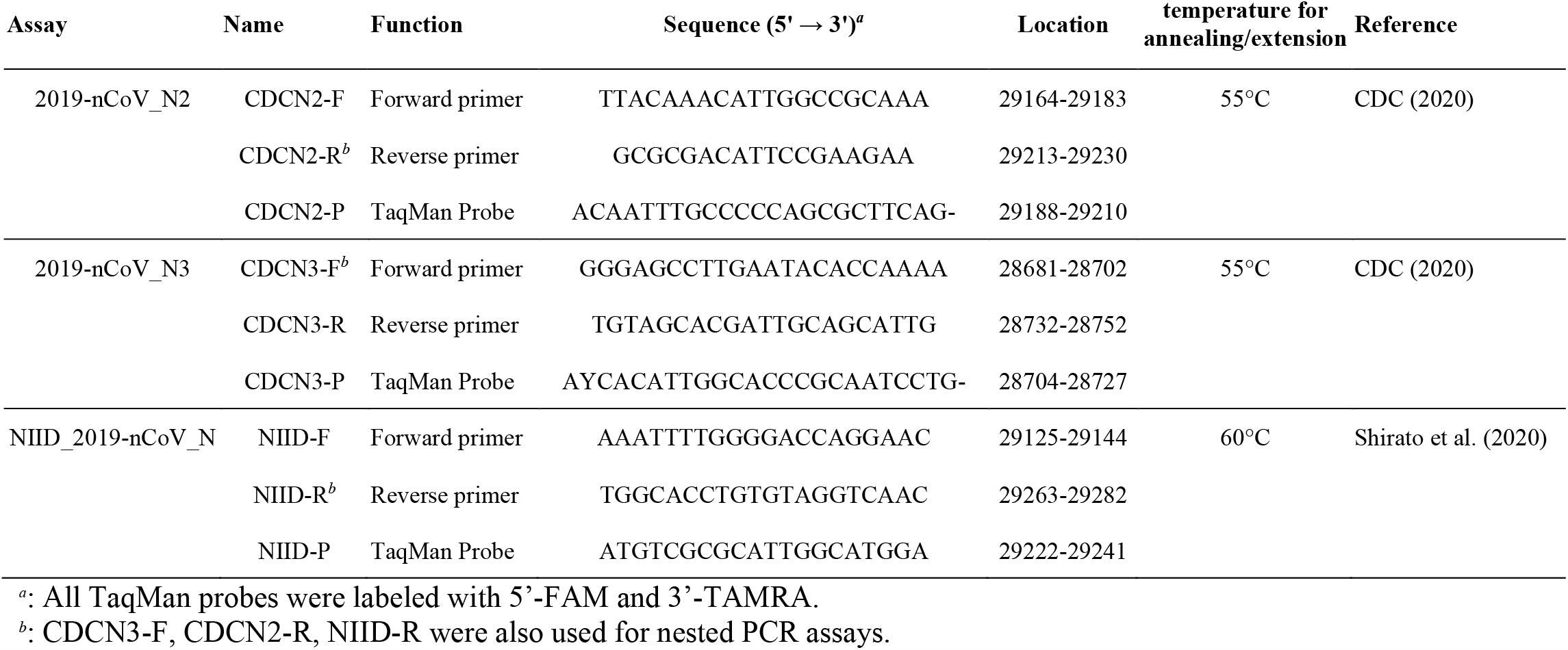
Primer and TaqMan probe sequences used for detection of SARS-CoV-2 in this study.

To obtain a calibration curve, a 10-fold serial dilution (concentrations ranged from 1.0×10^0^ to 1.0×10^6^ copies/reaction) of a standard DNA (plasmid DNA or oligo DNA) containing the target sequence was amplified. In each qPCR assay, 1.0×10^1^ copies/reaction of the standard DNA were positive at all times, indicating that the limit of the quantification values for each qPCR assay was below 1.0×10^1^ copies/reaction. If the resultant Ct value from a sample was corresponding to >1 copy/reaction, the sample was determined to be positive for SARS-CoV-2.

Absence of the positive signal in the no template control was confirmed in every qRT-PCR runs to exclude the potential contamination of the template into the reagents.

### 2.5 RT-nested PCR assays

The RT reaction was performed with a high-capacity cDNA reverse transcription kit (Life Technologies, Tokyo, Japan). The obtained cDNA was subjected to nested PCR assays for qualitative gene detection of SARS-CoV-2 (Fig. 1). The first round of PCR amplification was performed in 50 μL of a reaction mixture containing 10.0 μL of cDNA, 2×Premix Taq™ (TaKaRa), 400 nM each of CDCN3-F and NIID-R primers, which are also employed for CDCN3 and NIID qRT-PCR assays, respectively, and expected to amplify 600 bp gene fragment (Fig. 3 and Table 2), and nuclease-free water. The amplification was performed under the following thermal cycling conditions using a Thermal Cycler Dice (TaKaRa): initial denaturation and enzyme activation at 95°C for 10 min, followed by 40 cycles of amplification with denaturation at 95°C for 10 s, primer annealing at 55°C for 30 s, and an extension reaction at 72°C for 1 min and then a final extension at 72°C for 5 min. The first round PCR product was diluted with nuclease free water by 1,000-fold and then subjected to following PCR assays.

**Figure 3.**
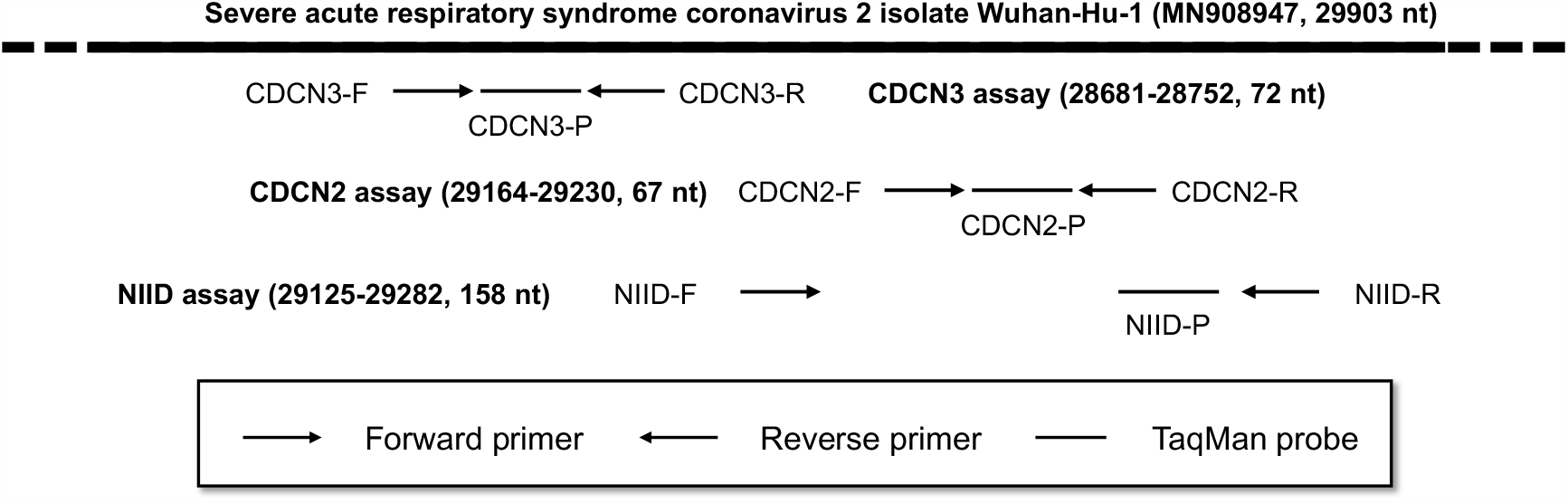
Location of primers and TaqMan probes employed in this study on a referential SARS-CoV-2 strain, Wuhan-Hu-1 (Gen Bank accession no: MN908947). These are localized on a relatively short fraction (600 nt) of N-gene. Therefore, CDCN3-F and NIID-R primers, for example, can be used as outer primers for nested assays.

For the second round of PCR, four different assays were conducted, one is a conventional PCR assay (semi-nested PCR assay) followed by gel electrophoresis and gene sequencing. Others are real-time PCR based on CDCN2, CDCN3, and NIID assays (nested-real time PCR assays). The semi-nested PCR assay was performed in a 20 μL reaction mixture containing 2 μL of the product of the first round of PCR amplification, 2×Premix Taq™ (Takara), and 400 nM each of CDCN3-F and CDCN2-R primers (Fig. 3 and Table 2), which are also employed for CDCN3 and CDCN2 qRT-PCR assays, respectively, and expected to amplify 550 bp gene fragment, and nuclease-free water. PCR amplification was performed under the following thermal cycling conditions using a Thermal Cycler Dice (TaKaRa): initial denaturation at 95°C for 10 min, followed by 40 cycles of amplification with denaturation at 95°C for 10 s, primer annealing at 55°C for 30 s, and extension reaction at 72°C for 30 sec and then a final extension at 72°C for 5 min. The semi nested PCR product was subjected to electrophoresis on a 1.5% agarose gel and visualized under a UV lamp after being stained with Gel Red™(FUJIFILM).

For further confirmation of the successful amplification by the first round PCR, the PCR product was subjected to CDCN2, CDCN3, and NIID assays (nested-real time PCR assays) with a qTOWER^3^ (Analytik Jena) (Fig. 3 and Table 2). The real time PCR was performed in a 20 μL reaction mixture containing 2 μL of the product of the first round of PCR amplification, 2×Premix Taq™ (TaKaRa), and 400 nM each of primers, 100 nM of a TaqMan probe, and nuclease-free water. PCR amplification was performed under the following thermal cycling conditions: initial denaturation at 95°C for 10 min, followed by 40 cycles of amplification with denaturation at 95°C for 10 s, annealing and extension at specific temperatures for each assay for 30 s (Table 2).

### 2.6 Determining efficiencies of virus concentration and RNA extraction-qRT-PCR

Indigenous F-phage in the sample before and after the concentration process was quantified by a plate counting assay employing *Salmonella Typhimurium* WG49 as a host strain, as described previously (Mooijman et al., 2002). The efficiency of the virus concentration process was estimated by comparing the F-phage concentration in the sample before the concentration process with that after the concentration process.

The efficiency of RNA extraction-qRT-PCR was estimated by comparing the observed gene concentration of the spiked MNV in nuclease-free water with that in the concentrate.

## 3. RESULTS

### 3.1 Efficiency of virus detection

The recovery efficiency of indigenous F-phage by the concentration process was 57% in geometric mean and was always above 10% (n = 27). RNA extraction-qRT-PCR efficiency determined with the spiked MNV was 66% in geometric mean and was always above 10% (n = 27). These indicate that the virus concentration and following molecular processes were conducted efficiency enough (Haramoto et al., 2018).

Presumptive virus detection efficiency, which can be estimated by multiplying these efficiencies, was 38% in geometric mean and below 10% for 5 out of 27 samples (3.7% was the lowest).

### 3.2 Detection of SARS-CoV-2 by qRT-PCR assays

In this study, three qRT-PCR assays, CDCN2,CDCN3 and NIID assays, were separately applied. A sample collected at WTTP D on April 3 was positive for SARS-CoV-2 using all the assays with an observed concentration of around 3.0×10^4^ copies/L (2.1×10^4^ – 4.4×10^4^ copies/L) (Fig. 2 and Table 3). The presumptive virus detection efficiency in the sample was relatively high (estimated as 130%). In addition to the sample, CDCN2, CDCN3, and NIID assays showed positive signals in 1 (WWTP B on April 16), 4 (WWTP A on March 19, WWTP B on March 26 and April 23, and WWTP C on April 21), and 1 (WWTP D on April 24) samples. In total, 7 samples were determined to be positive for SARS-CoV-2 by at least one of the assays with observed concentration of 1.2×10^4^ – 4.4×10^4^ copies/L. The detection frequency seems to be higher when the number of total confirmed SARS-CoV-2 cases in 100,000 peoples became above 10 in each prefecture (Fig. 2 and Table 3). The SARS-CoV-2 detection frequency was 15% (3 positives out of 20 samples) before the number became >10, whereas it reached 57% (4 positives out of 7 samples) after the number became >10.

**TABLE 3.** Summary of samples positive for SARS-CoV-2 by qRT-PCR assays.

| Date | WWTP<sup>a</sup> | Total confirmed cases<br>in 100,000 peoples |  | Efficiency of detection process |  |  | qRT-PCR |  |  |  | RT-nested PCR |  |  |
| --- | --- | --- | --- | --- | --- | --- | --- | --- | --- | --- | --- | --- | --- |
|  |  | Ishikawa | Toyama | Concentration | RNA extraction-qRT-PCR | Total | CDCN2 | CDCN3 | NIID | semi-nested | CDCN2 | CDCN3 | NIID |
|  |  |  |  | % | % | % | copies/L |  |  |  | Ct | Ct | Ct |
| March 19 | A | 0.4 | 0.0 | 263 | 72 | 190 | Neg.<sup>b</sup> | 2.8×10<sup>4</sup> | Neg. | Neg. | Neg. | 35.2 | Neg. |
| March 27 | B | 0.5 | 0.0 | 76 | 50 | 38 | Neg. | 1.7×10<sup>4</sup> | Neg. | Neg. | Neg. | 36.2 | Neg. |
| April 3 | D | 1.4 | 0.8 | 125 | 102 | 130 | 4.4×10<sup>4</sup> | 1.2×10<sup>4</sup> | 2.1×10<sup>4</sup> | Neg. | Neg. | 29.9 | Neg. |
| April 16 | B | 12.2 | 5.1 | 10 | 49 | 4.9 | 1.5×10<sup>4</sup> | Neg. | Neg. | Pos. | Neg. | Neg. | Neg. |
| April 21 | C | 15.7 | 10.9 | 87 | 41 | 36 | Neg. | 1.5×10<sup>4</sup> | Neg. | Pos. | Neg. | 32.1 | Neg. |
| April 23 | B | 17.4 | 11.0 | 36 | 50 | 18 | Neg. | 1.2×10<sup>4</sup> | Neg. | Pos. | Neg. | 31.1 | Neg. |
| April 24 | D | 18.9 | 14.3 | 51 | 31 | 16 | Neg. | Neg. | 1.4×10<sup>4</sup> | Pos. | Neg. | 30.3 | Neg. |

<sup>a</sup>: WWTPs A-C and D are located in Ishikawa and Toyama prefecture, respectively.
<sup>b</sup>: Neg.: Negative; Pos.: Positive.

### 3.3 Detection of SARS-CoV-2 by RT-nested PCR assays and nucleotide sequencing

The qRT-PCR assays used in this study target a 600 bp fraction of gene coding N-protein. Then, nested PCR assays could be performed using the primers used in the qRT-PCR assays for further confirmation of the positive results obtained by the qRT-PCR assays. A semi-nested PCR assay to amplify 550 bp of a gene fragment showed a band of around 600 bp size in four out of seven samples, which were collected during April (Table 3 and Fig. 4). However, direct Sanger sequencing could not confirm if the amplicon was obtained from SARS-CoV-2 or not, because of too many heterozygous peaks. Application of real-time PCR to the first round PCR products showed ambiguous results. CDCN2 and NIID assay did not show any positive results, while CDCN3 assay showed positive signal from six out of the seven samples. Among them, samples collected during March resulted in relatively high (>35) Ct values, while those collected during April resulted in lower (around 30) Ct values.

**Figure 4.**
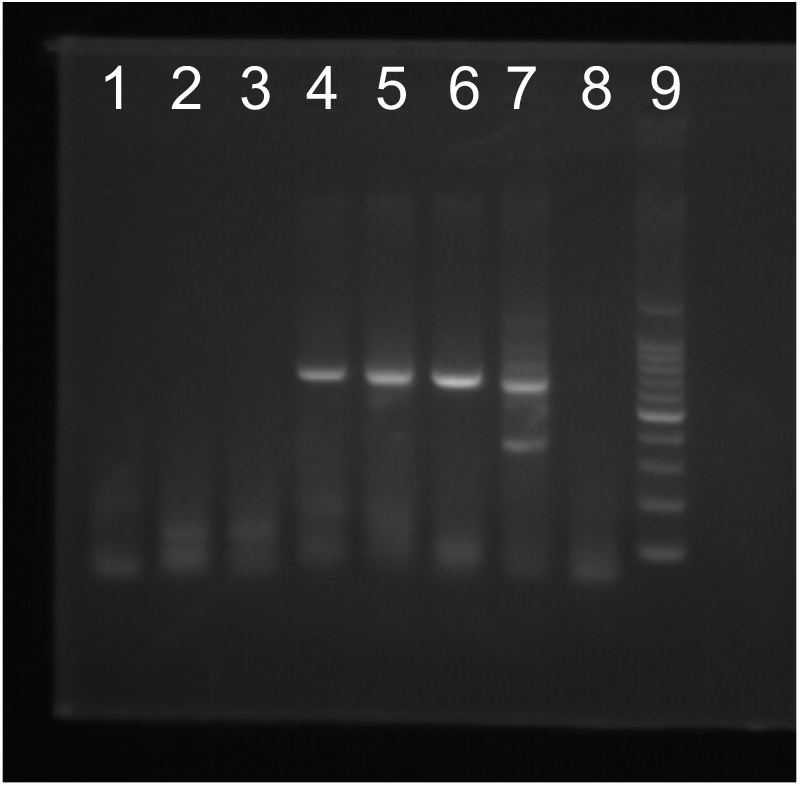
Detection of SARS-CoV-2 in wastewater samples by the RT-semi-nested PCR assay, which is expected to result in a 550 bp of amplicon. Lanes 1-3: Samples collected at WWTPs A, B, and D on March 19, 26, and April 3, which did not show the positive band. Lanes 4-7: Samples collected at WWTPs A, B, C, B, on April 16, 21, 23, and 24, which showed the positive band. Lane 8: Non target control. Lane 9: 100-bp ladder.

## 4. DISCUSSION

Most of previous studies investigated pathogenic and indicator viruses targeted non-enveloped viruses (Haramoto et al., 2018). Consequently, retrospective virus concentration methods are optimized for non-enveloped viruses and may not be effective to enveloped virus like SARS-CoV-2. For instance, adsorption-elution methods using electropositive or electronegative microfilter and glass wool are surely effective to non-enveloped viruses (Cashdollar et al., 2013) but resulted in inefficient recovery efficiency (consistently less than or around 10%) for at least some of enveloped viruses (Haramoto et al., 2009; Honjo et al., 2010; Deboosere et al., 2011; Francy et al., 2013; Blanco et al., 2019). A size excusive method using ultrafiltration is also widely applied for virus monitoring (Cashdollar et al., 2013). Even though its efficacy in concentrating enveloped viruses has not been fully elucidated yet, some researchers have been succeeded in detecting enveloped viruses, influenza-A virus and SARS-CoV-2 (Heijnen and Medema, 2011; Medema et al., 2020; Ahmed et al., 2020; Nemudryi et al., 2020).

One of the drawbacks of the ultrafiltration-based method is the cost of the ultrafiltration unit. For example, Centricon plus-70, which can process up to 70 mL of a sample at once, costs approximately 50 USD per unit, and it might not be suitable for frequent use. On the contrary, PEG precipitation is one of the most cost-effective method for concentrating virus in a low volume (around 100 mL) of water sample (Lewis and Metcalf, 1988). The technique is effective not only for viruses but also for variety of proteins (Atha and Ingham, 1981). Therefore, it might be effective for SARS-CoV-2, even though only a limited number of studies have applied the technique to enveloped viruses, including SARS-CoV-2, in water (Honjo et al., 2010; Deboosere et al., 2011; Wu et al., 2020). For PEG precipitation, several conditions, i.e., concentrations of PEG and NaCl, have been applied (Jones and Johns, 2009). Among them, we selected relatively high concentrations of PEG and NaCl (10% and 1.0 M), in accordance with Jones and Johns (2009). As expected, the method revealed a sufficiently high and stable recovery efficiency of indigenous F-phages (57% in geometric mean), indicating that SARS-CoV-2 was also recovered efficiently. An efficient way for recovering SARS-CoV-2 in wastewater and an appropriate recovery control to ascertain the efficient recovery of SARS-CoV-2 should be found in future.

Several molecular assays have been designed for detection of SARS-CoV-2 (CDC2020; Shirato 2020; Corman et al., 2020; Chu et al., 2020). Although some studies revealed effectiveness of these assays in comparative manner (Jung et al., 2020), a gold standard method has not been determined yet. Especially, wastewater contains variety of microbes and free nucleic acids, which might lead to false positive due to non-specific amplification. Therefore, most studies on SARS-CoV-2 in wastewater samples applied multiple RT-PCR assays, gel electrophoresis, and amplicon sequencing in attempts to improve reliability (Ahmed et al., 2020; Medema et al., 2020; Nemudryi et al., 2020; Wu et al., 2020; Wurtzet et al., 2020). In this study, three qRT-PCR assays and additional RT-nested PCR assays, to confirm the positive results obtained by the qRT-PCR assays, were applied. Among seven samples positive by at least one of the qRT-PCR assays, one collected at WWTP D on April 3 were consistently positive by all three qRT-PCR assays, while other six samples were positive by one of the assays. Low concentrations of SARS-CoV-2 gene in the concentrates can be one possible reason for the ambiguous results. The observed concentration was at most 8 copies/reaction. In this case, the target gene is occasionally absent in the analyte. It is also possible that some of the positive signals were generated by non-specific amplicons or that some assays failed to amplify specific SARS-CoV-2 strains.

Then, RT-nested PCR assays sharing primers with the qRT-PCR assays were conducted to ascertain the positives obtained by the qRT-PCR assays. Most of the qRT-PCR positive samples again showed positive results by at least one of the RT-nested PCR assays, indicating that the positive signals obtained by the qRT-PCR assays are reliable. Some previous studies revealed that CDCN2 assay seems to be less efficient in detecting SARS-CoV-2 in wastewater samples than CDCN1 and CDCN3 assays (Medema et al., 2020; Wu et al., 2020; Nemudryi et al., 2020; Randazzo et al., 2020). Accordingly, CDCN2 assay, as well as NIID assay, seem to be inefficient in this study, showed no positive results after the first round PCR. Similarly, the first round PCR followed by CDCN3 assay seems to be better than that followed by the semi nested PCR assay, in view of the number of positives. These suggest that primers and TaqMan probe used in CDCN2 and NIID assays might contain mismatches with SARS-CoV-2 strains in wastewater. Notably, one sample (at WWTP B on April 16) failed to confirm the presence of SARS-CoV-2 by the second round CDCN3 assay. This sample was also negative by qRT-PCR with CDCN3 assay. Considering that the sample was positive by the semi nested assay, the first round PCR was successfully conducted. This implies that CDCN3 assay may also not be effective in detecting all the circulating SARS-CoV-2 strains.

Detection frequency of SARS-CoV-2 in our wastewater samples seemed to be in agreement with the numbers of confirmed cases in both prefectures. The detection frequency seems to be higher when it becomes above 10 in 100,000 peoples (Fig. 2 and Table 3) Besides, no positive result was obtained in the samples collected in Toyama prefecture during March, when no confirmed SARS-CoV-2 infection cases were reported. Our results also suggest that SARS-CoV-2 in wastewater sample could be identified even when the number of cases per 100,000 peoples is below 1.0, although the detection frequency becomes low (Fig. 2 and Table 3). One SARS-CoV-2 positive sample during March was collected at WWTP A. A relatively low number of populations served by the WWTP (31,510 peoples) might be attributed to the positive result. Smaller WWTP would have a limited dilution effect and can be more sensitive to the presence of the number of infected individuals. Previous studies have suggested that the concentration of SARS-CoV-2 in wastewater is affected by number of peoples infected in the catchment area (Medema et al., 2020; Ahmed et al., 2020; Wurtzer et al., 2020b). However, there seems to be a discrepancy in the observed relationships between the concentration in wastewater and the number of infected people. Ahmed et al. (2020) could not identify SARS-CoV-2 in a wastewater sample in Queensland until the number of the confirmed cases reached up to around 100 cases per 100,000 peoples. In addition, the observed concentration was below the limit of quantification at that time. In contrast, Wurtzer et al. (2020b) repeatedly obtained positive results in Paris even when the number of the confirmed cases was below 1 case per 100,000 peoples, in accordance with our observation. One of the reasons for this discrepancy might be due to the difference in methods applied; collecting composite samples or grab samples and ways of sample concentration, nucleic acid extraction, and qRT-PCR. Besides, the number of the confirmed cases does not necessarily reflect the actual prevalence of the infection at the time point, which can be better correlated to the concentration in the wastewater. In Japan, it takes a few days to confirm the case by the clinical surveillance since the symptom first appeared or the one become infected. There should be asymptomatic cases, which were not confirmed. In the context of WBE, data on the relationship between the number of infection cases and concentration in wastewater needs to be accumulated further.

## Data Availability

All the data are available in the manuscript

## ACKNOWLEDGEMENTS

This work was supported by Hiramoto-gumi Inc., the civil construction company in Japan.

